# Developing Standards for Rapid Evaluation and Appraisal Methods (STREAM): an e-Delphi consensus study

**DOI:** 10.1101/2023.12.11.23299803

**Authors:** Sigrún Eyrúnardóttir Clark, Norha Vera San Juan, Cecilia Vindrola-Padros

## Abstract

Timeliness is key to influencing the utility of evaluation and research findings and has given rise to a range of rapid evaluation and appraisal approaches. However, issues in the design, implementation and transparency in their reporting has led to concerns around their rigour and validity. To address this, we have developed the Standards for Rapid Evaluation and Appraisal Methods (STREAM). We followed a four-stage consensus process, starting with a (1) steering group consultation; (2) three-stage e-Delphi study; (3) stakeholder consensus workshop; and (4) piloting exercise. The stakeholders invited to participate in the consensus process had experience in conducting, being part of, or commissioning rapid evaluations or appraisals. As a result, 38 standards were developed with the purpose of guiding the design and implementation of rapid evaluations and appraisals and supporting the reporting of methods used. We suggest strengthening STREAM by testing it in alternative contexts to assess its generalisability.

## 1. Introduction

Timeliness has been highlighted as a key factor influencing the utility of evaluation and research findings such as in response to humanitarian crises or the evaluation of new or changing services (Heather Nunns, 2009). A wide range of rapid evaluation, assessment and appraisal approaches have been developed to make findings available when they are most needed. These approaches are characterised by the short duration of evaluation or research, use of multiple methods for data collection, teams of researchers or evaluators, formative study designs where findings are fed back while the data collection is ongoing, and the development of actionable findings (adequate for purpose) to inform changes in policy and/or practice (Anker et al., 1993; Beebe, 1995; James Beebe, 2014; McNall & Foster-Fishman, 2007). Rapid appraisals are used when findings are delivered in time and resource-limited environments (Vindrola-Padros, 2021). Whilst rapid evaluations are often used to produce evidence on services or programs that can inform decision making on their delivery (Vindrola-Padros, 2021).

Challenges exist in the design and implementation of rapid approaches. Researchers and evaluators often face tensions between the breadth and depth of data included in studies, which raises questions regarding validity (Manderson & Aaby, 1992a, 1992b; Vindrola-Padros & Vindrola-Padros, 2018). For instance, short-term data collection periods might not allow researchers or evaluators to capture changes over time, understand all relevant socio-cultural factors at stake or document conflicts and contradictions in findings, thus potentially leading to unfounded interpretations and conclusions (Bentley et al., 1988; Harris et al., 1997). Additionally, as rapid approaches often rely on team-based methods, variability between researchers and evaluators may influence the reliability of the data (Bentley et al., 1988; Manderson & Aaby, 1992a). Shorter fieldwork periods also raise questions in relation to the representativeness of samples as evaluators and researchers may need to rely on the participants who are most accessible, losing diversity in experiences and points of view (Bentley et al., 1988; Harris et al., 1997; Manderson & Aaby, 1992a; Utarini et al., 2001). Researchers and evaluators might not have time to follow-up with participants to cross-check information or explore additional topics. Periods of data analysis might need to be compressed, affording little time for critical reflection (Pink & Morgan, 2013; Utarini et al., 2001). Another common issue lies in the lack of transparency in the methods used and changes made throughout these types of studies (Johnson & Vindrola-Padros, 2017; Vindrola-Padros & Vindrola-Padros, 2018).

One way to improve the transparency and completeness of reporting and increase the quality of studies is through the development of reporting standards (Ogrinc et al., 2016; Tong et al., 2007). Standards can provide evaluators and researchers with guidance to improve the quality and validity of their rapid studies. Standards can also be used to promote clear and detailed reporting of study methods, adaptations and limitations. There are currently no published standards or guidelines for rapid evaluations and appraisals.

The aim of this study was to develop the first Standards for Rapid Evaluation and Appraisal Methods (STREAM) to be used in broad contexts (not just restricted to health), through the use of a series of consensus building steps. This included an e-Delphi study which is a method used to arrive at a group consensus or decision by surveying a panel of experts in the field of rapid evaluation and appraisal methods using an electronic platform over multiple rounds (Akins et al., 2005; Hasson et al., 2000; Howarth et al., 2021; McKenna, 1994; Moher et al., 2010). This was followed by conducting collaborative stakeholder workshops to improve the clarity of statements; and finally facilitating a piloting exercise to understand the validity of STREAM in practice. The research team previously conducted a systematic review to identify the methods that had been used to ensure rigour, transparency and validity in rapid evaluation, appraisal and assessment methods (S. Clark et al., 2022). The findings from the systematic review were used to guide the development of a list of items to include in the first round of the e-Delphi consensus process.

## 2. Methods

### 2.1 Protocols and ethical approvals

A protocol was developed and published on the Open Science Framework (OSF) network as a means to guide the project (S. E. Clark et al., 2022). This project was also registered on the Enhancing the QUAlity and Transparency Of health Research (EQUATOR) network (EQUATOR Network, 2022). Established guidance by Moher et al. for the development of health research reporting guidelines was followed throughout this study (Moher et al., 2010). Ethical approval was received from the UCL Research Ethics Committee under the project ID: 23555/001.

### 2.2 Systematic review

The systematic review findings were used to identify the methods that had been used to ensure rapidity of studies whilst maintaining transparency and rigour (S. Clark et al., 2022). The research team reviewed the methods, and created an initial list of statements that could be used to guide the methods chosen for future rapid studies.

### 2.3 Steering group

A steering group was established consisting of a lived experience researcher in the field of Primary Care and Public Health; a researcher in the field of Psychology and Neuroscience; a Health Policy researcher; and a global health researcher. The four steering group members were able to provide feedback on the draft items that were identified from the systematic review to include in the first round of the e-Delphi study, they were also able to suggest items for inclusion that had not been identified from the review. The group were also able to circulate invitations to the study within their networks.

### 2.4 Delphi Study

#### 2.4.1 Creation of the survey

The Delphi survey was created using the online platform Welphi (Welphi, 2023). The survey itself consisted of a consent page informing that participation was voluntary, how data will be used and how long it will be stored for. This was followed by demographic questions that asked participants about their field of work; duration of experience in the field; whether they were a carer or individual with lived experience; where the participants were based and the locations of their research.

The survey then went on to list the first round of Delphi items in a ranking exercise. Next to each statement was a Likert scale asking respondents to rate how relevant they thought each statement was on a scale of 1 as irrelevant to 4 as relevant. There was also an option for ‘Don’t know’ and an option for participants to write a comment next to each statement, should they think the statement could be reworded, or if they had any general comments.

The ranking exercise in the survey was then followed by a free text question, which allowed participants to suggest other additional items to include in the standards or to share any general comments. This feature allowed for an open round of data collection.

#### 2.4.2 Rounds of the survey and thresholds

The plan was to conduct three rounds of the survey, with thresholds of 70% and 15% for each round. Meaning that if 70% or more of participants voted an item as relevant, and if 15% or less of participants voted an item as irrelevant, that item would have reached consensus for inclusion in the next round. If 70% or more of the participants voted an item as irrelevant, and if 15% or less of participants voted an item as relevant that item would have reached consensus for exclusion in the next round. If an item was neither voted by 70% or more of participants as relevant or irrelevant it would also be included in the next round. Although a set level of consensus does not exist for the Delphi method, these thresholds have been used previously in consensus studies (Hasson et al., 2000; Williamson et al., 2012). The open round of data collection and the ability to comment on the wording of statements was only facilitated in the first round of the survey.

Once the first round of the survey was developed, it was piloted among six members of the wider research team. Feedback was shared on the clarity of email invitations, the functionality and accessibility of the platform and survey, and if there were any errors in the format or wording of the survey.

#### 2.4.3 Sampling and eligibility criteria of Delphi participants

There were 283 potential participants that were purposively sampled and invited to participate. These participants were identified based on recommendations from the research team; recommendations from the steering group; from authoring or editing publications or reports in the field of rapid evaluation and appraisals; and as recommendations from participants who had already been invited to participate (a form of snowball sampling). The inclusion criteria to be selected as a participant was that they needed to have experience in conducting, participating, reviewing or using findings from rapid studies. The target sample size for the Delphi study was 50-80 participants, taking into consideration attrition (the likelihood of losing participants with each round of the survey).

#### 2.4.4 Data collection

It was planned that each round of the survey would remain open for two weeks after invitations had been shared. The survey invitations for the first round of the survey were shared with potential participants on the 24^th^ February 2023, however due to initial poor response rates, the survey remained open for four weeks. The subsequent second and third rounds of the survey took place between 31 March – 21 April, and 26 April – 12 May respectively.

#### 2.4.5 Data analysis

After each round of the survey, the results were exported into a Microsoft Excel file, which included a summary produced by the Welphi platform of the basic statistics -the percentages of consensus for each statement. This allowed the research team to identify statements that had reached the threshold for inclusion into the next round of the Delphi, or for final inclusion in the Delphi statements. These decisions also considered the free text comments made by participants in the first round of the survey.

The research team were able to descriptively analyse these comments, and agree any changes that should be made to the wording of statements, or any additional items to include in the second round of the Delphi study.

### 2.5 Stakeholder workshop

Following the analysis of the responses from the final round of the Delphi survey, the research team collated the list of the final items to develop STREAM. These final items were then shared with stakeholders at a collaborative workshop in June 2023. These stakeholders were individuals who had experience or interest in conducting rapid appraisals and rapid evaluations, or had acted as commissioners of rapid evaluations and appraisals.

The stakeholders were asked to provide feedback on the clarity and order of the statements within STREAM. They were also asked to look at three statements in depth that had not reached 70% consensus in the final round of the Delphi study for inclusion or exclusion. Field notes were taken by the research team to capture the discussion points and update STREAM.

### 2.6 Pilot scenarios

The updated statements were then used in a piloting exercise. A member of the broader research team used STREAM to guide the reporting of a rapid evaluation study looking into student nurse experiences of a pilot program rolled out in five inner city hospitals in the UK. The study had been conducted between August and November 2022, the researcher then reviewed and used STREAM after these dates to guide the development of a publication for submission to a journal summarising the methods they had used and their findings.

Following the use of STREAM, the researcher who had participated in the piloting exercise shared feedback on the applicability of each statement in their context. This feedback was shared during a one-to-one discussion, and captured in the form of field notes. The feedback was used to make changes in the final version of STREAM.

## 3. Results

### 3.1 Drafting items based on the systematic review findings and steering group feedback

Reviewing the methods identified from the systematic review led to the development of 32 draft items for the Delphi (see Appendix A). The research team grouped these items into seven categories: study design; research team; data collection; data analysis; result interpretation; dissemination; governance and accountability.

All four members of the steering group provided feedback on the draft Delphi items. Their feedback ranged from making changes to the general structure of the statements; changes to the study design statements; changes to the research team statements; changes to the result interpretation, dissemination and governance and accountability statements. These can be found summarised in Table 1 below. The draft Delphi items were updated to reflect the feedback which resulted in a total of 36 statements that fed into the first round of the e-Delphi survey (see Appendix B).

**Table 1.** Summary of feedback from the steering group.

| Themes of feedback | Feedback |
| --- | --- |
| Layout and structure of the statements | • Ensure there is no overlap between the items as this would make it difficult to use the statements as a critical appraisal tool.<br>• Change the order of the statements and move them between the seven categories listed above, for better flow and readability.<br>• Use clearer phrases and terms that all would be able to understand, such as describing what patient, public involvement is. |
| The study design statements | • Include a statement on how the data is likely or planned to be used in the study design section. |
| The research team statements | • Include a statement on the geographical location of researchers in comparison to the research site as those who are located in the field are able to respond and evaluate the situation much quicker.<br>• Include a statement on whether lived experience researchers were included within the research team. |
| The interpretation, dissemination and governance statements | • Include a statement to consider if recommendations were made as a result of the study findings. |
|  | It is difficult to report on how the study findings were used as this is often not determined by the evaluators or appraisers, more so the commissioners.<br>Include a statement on the funding source. |

**Table 2.** Characteristics of the participants included in the first round of the study (n=60)

| Characteristics | Number of participants (percentage) |
| --- | --- |
| Field of work: |  |
| Health | 53 (88.3%) |
| Lived experience researcher | 1 (1.7%) |
| Education researcher | 1 (1.7%) |
| Evaluation professional (no mention of health) | 5 (8.3%) |
| Years of experience: |  |
| ≤5 years | 8 (13.3%) |
| > 5 years and <20 years | 32 (53.3%) |
| ≥ 20 years | 20 (33.3%) |
| Carer or individual with lived experience: |  |
| Yes | 15 (25.0%) |
| No | 42 (70.0%) |
| Unsure | 3 (5.0%) |
| Country individual is based in: |  |
| Australia | 1 (1.7%) |
| Mexico | 2 (3.3%) |
| New Zealand | 2 (3.3%) |
| Republic of Ireland | 3 (5.0%) |
| Republic of Liberia | 1 (1.7%) |
| Spain | 1 (1.7%) |
| Switzerland | 1 (1.7%) |
| United Kingdom | 34 (56.6%) |
| United States of America | 15 (25.0%) |
| Location where work is conducted: |  |
| Outside of the country participants are based in | 21 (35.0%) |
| Within the country participants are based in | 39 (65.0%) |

### 3.2 Consensus from the Delphi study

#### 3.3.1 Delphi sample size and characteristics

From the 283 participants invited to take part in the study, 60 (21.2%) participated in the first round, 49 (17.3%) in the second, and 47 (16.6%) completed all three rounds of the Delphi survey. This means the attrition rate across the three rounds was 21.7%.

As demonstrated in the table above, the majority of participants worked in the field of health which encompassed many sub fields such as health psychology, medical anthropology, health services research, public health, health policy research, among others. The majority of participants were based in the UK, followed by the USA, and 35% of participants conducted their work or research outside of the country they resided in. A quarter of the participants either had lived experience in terms of using health care services, or experiencing health conditions or had cared for someone else with a health condition.

#### 3.2.2 Rates of consensus and comments on the statements from the first round of the Delphi

There were between 58-60 responses to the ranking of statements following the first round of the Delphi. Consensus was reached on 29 of the 36 items, whereby 70% or more of the participants ranked the statements as three or four on the Likert scale which represented relevance for inclusion in the standards. However, nine of the statements had between 16-30% of participants voting them as irrelevant (one or two on the Likert scale), meaning consensus was not achieved on these statements. There were seven items that also didn’t reach consensus, whereby less than 70% of the participants thought the items were relevant, however consensus was not reached for excluding these items, as across the items only 28-49% of participants thought that they were irrelevant. These items were therefore included in the next round of the Delphi, along with the items that reached consensus on inclusion.

Prior to the second round of the Delphi the statements were updated based on the qualitative feedback from the open round of voting, a summary of the types of suggested amends can be found in Table 3 below.

**Table 3.** Summary of feedback from the first round of the Delphi: open voting round.

| Themes of feedback | Feedback |
| --- | --- |
| General | • Re-ordering statements for a better flow and readability.<br>• Merging statements that seem repetitive and separating a statement that consists of too much information or feasible actions.<br>• Include 'if appropriate' to some statements, to make it clearer that not all are mandatory depending on the contexts in which the statements are used.<br>• It may be important to have a short form and longer form of the standards, one that can be generalised to all contexts and then other shorter forms that are more specific to different contexts.<br>• A greater focus of participation, diversity and inclusion.<br>• One participant thought developing a checklist may prevent creativity and flexibility in research. |
| Study design | • Describing any preliminary research or what shaped decisions around study design may be too onerous and take away from important time in implementing rapid studies.<br>• Include further information on what should be included in study protocols such as patient public involvement and engagement planned.<br>• Include a statement on the study duration including the data collection and analysis periods.<br>• Report on if any changes were made to the study design during data collection. |
|  | • A contrasting comment was that protocolising everything in qualitative studies could be limiting, whilst another participant thought protocols may not be so relevant to evaluation studies. |
| The research team | • Highlight if any changes were made to the research team over time.<br>• Make it clearer that you want to understand the researcher's relationship and proximity to the research site in terms of whether they are local to the research site or whether they are conducting research virtually. However this may be less relevant if evaluations were conducted domestically.<br>• Some participants didn't think the research team size, expertise or roles and responsibilities should determine the quality of the study.<br>• Some participants didn't think including a statement on training was necessary, some found it patronising whilst others thought it should be specified to training in rapid methods only. |
| Data collection and analysis | • Provide further guidance on how to ensure cultural validity and conceptual equivalence in any translations.<br>• Make is clearer that the focus should be on maintaining consistency in the methods used.<br>• Incorporate the term approach rather than tools, as not all researchers will use specific tools for data collection, rather they will use a general approach.<br>• Provide further guidance on how to triangulate data. |
| Result interpretation | • Provide more information on what member checking is.<br>• Some found member checking problematic.<br>• Some thought reflexive practice as a statement should be optional as it may not be relevant in all study contexts and could be time consuming to implement.<br>• Some thought that linking findings to existing published literature and generalisability or transferability with other populations was more relevant to research projects and academia than evaluations. |
| Dissemination | • Provide further information on what dissemination could look like for different audiences.<br>• It may not be possible for some to report on how findings were used as commissioned researchers or evaluators, as the information is not shared back by the commissioners.<br>• Include a statement on data sharing and access to raw data. |
| Governance and accountability | • Make it clearer that regulatory approvals encompass ethical approvals too. |

As a result of incorporating the amends from the qualitative feedback, 36 items were included in the second round of the Delphi. These can be found in Appendix C, these included statements that had been updated based on modifications to wording, some statements had been split into separate statements, whilst others that were very similar had been combined. No statements were categorically removed based on comments, as consensus had not been achieved on the voting to remove any statements. Some of the additional comments were not incorporated in the updated statements but will instead be taken into consideration when developing a future explanation and elaboration document.

#### 3.2.3 Rates of consensus on the statements from the second round of the Delphi

Following the second round of voting, there were between 46-49 responses to the ranking of statements. There were 32 out of the 36 statements that had been ranked as relevant (three or four on the Likert scale) by 70% or more of the participants, 11 of the statements did however have between 15-29% of participants voting them as irrelevant (one or two on the Likert scale), meaning consensus was not achieved on these statements. There were four statements that also didn’t reach consensus, whereby less than 70% of the participants thought the items were relevant, however consensus was not reached for excluding these items, as across the items only 31-57% of participants thought that they were irrelevant. All of the items were therefore included in the third round of the survey again.

#### 3.2.4 Rates of consensus on the statements from the third round of the Delphi

After the third and final round of voting, there were 44-47 responses to the ranking of the statements. There were 33 out of the 36 statements that had been ranked as relevant (three or four on the Likert scale) by 70% or more of the participants, 10 of the statements did however have between 17-26% of participants voting them as irrelevant (one or two on the Likert scale), meaning consensus was not achieved in relation to these statements. There were three statements that also didn’t reach consensus, whereby less than 70% of the participants thought the items were relevant, however consensus was not reached for excluding these items, as across the items only 32-64% of participants thought that they were irrelevant.

### 3.3 Feedback from the stakeholder workshop

All of the items from the final round of the Delphi study were then shared at a stakeholder workshop, with special attention paid to the three items that had not reached consensus for inclusion across 70% or more of the participants (listed as quotations below).

*“Provide a clear description of the research team size (including any changes over time).”*

*“Indicate if team members received any training in rapid research methods.”*

*“Describe the roles and responsibilities of team members in this project and why the team was designed in this way.”*

The full list of items that the stakeholders reviewed can be found in Appendix C. A summary of their feedback can be found in Table 4 below. All of the stakeholders agreed that the three statements listed above should remain within the standards. They suggested one modification, that the examples of types of training could be included in the explanation and elaboration document.

**Table 4.** Summary of feedback from the stakeholder workshop.

| Themes of feedback | Feedback |
| --- | --- |
| General | • Update the numbering format of statements to letters under each numbered subheading.<br>• Emphasise more clearly that the statements are relevant to evaluation too, not just research.<br>• Some statements may be more relevant to a rapid appraisal versus a rapid evaluation, make this clearer in the explanation and elaboration document.<br>• Update the ordering and wording of statements for better readability. |
| Study design | • Include programme theories and theories of change within the theoretical frameworks or models that could guide study design.<br>• Provide examples within the explanation and elaboration document about the types of changes to protocols that would be important to report on.<br>• Don't limit reasons for not including certain sampling groups to time pressures. |
| Research and evaluation team | • Make it clearer about what is meant by the researcher/evaluator relationship and proximity to the research/evaluation site.<br>• Share examples of types of training that may be relevant to team members in the explanation and elaboration document. |
| Data collection and analysis | • Provide further guidance on how to ensure cultural validity and conceptual equivalence in any translations in the explanation and elaboration document. |
| Result interpretation | • Comparing generalisability of findings is not a common approach to take in evaluations, so remove this from the statement.<br>• Often evaluations do not share limitations of the study, instead they focus on gaps identified from the evaluation, so update the statement to include this. |
| Dissemination | • Dissemination and impact are two separate areas, so re-categorise the statements to reflect this. |

### 3.4 Findings from the piloting exercises

The team member responsible for piloting the standards in the reporting of their rapid evaluation for submission to a publication, was able to implement either partially or fully the majority (32 of 38) of the statements in their reporting. The reasons for not being able to use all or some of the statements are listed below.

- Any statements related to the planned deliverables were irrelevant for this project, as the evaluation team did not have transparency on how their findings had been used, the commissioner of the evaluation project did not share this information with them.
- Patient and Public Involvement and Engagement and community participation was not considered within this study as their sample included primarily healthcare professionals.
- No translation was required as all study participants spoke English, the same language as the evaluation team and commissioner.
- Member checking was not an approach used for this study.

The statements above may have been incorporated into the reporting of this project if the evaluation team had used the standards to guide their study design instead of just reviewing the standards following completion of their project for reporting.

There were some statements as a whole or subsections within statements that this piloting exercise did not identify as necessary to include in the reporting of their project for publication. This was because they thought it was irrelevant to the requirements of their journal of interest, and would bring their publication over the word count. These areas have been listed below:

- The whole study duration or specifically the data analysis period was not reported on, they did however report on the data collection period.
- The levels of experience of all team members and their backgrounds. They did however report on the professional backgrounds on a sub sample of the team members.
- They did not share whether researchers/ evaluators reflected on how their background and experiences may have affected their data collection, analysis and interpretation.
- They did not confirm if it was possible to access the raw data from the study on request, especially as they did not know if this was appropriate without permission from the commissioner of their study.

No amends were made to the statements following this piloting exercise, but a review plan has been developed to regularly update STREAM after it is applied in other settings.

### 3.5 The standards for Rapid Evaluation and Appraisal Methods (STREAM)

As a result of the consensus process and steps listed above, STREAM was developed. The current list can be found in Table 5 below. An explanation and elaboration document will be developed, to provide further information on each statement listed in the table.

**Table 5.** The Standards for Rapid Evaluation and Appraisal Methods (STREAM): July 2023.

|  |
| --- |
| **1. Study design** |
| a) Define the purpose, aim or research/ evaluation questions and planned deliverables guiding the study. |
| b) Provide a clear description of the intervention, programme or service being evaluated. |
| c) Describe any preliminary research, scoping studies or piloting methods to inform the study design. |
| d) Describe any theoretical frameworks or models used to guide the study design (or programme theories or theories of change in the case of rapid evaluations). |
| e) Indicate any relevant reporting guidelines used throughout the study. |
| f) If Patient and Public Involvement and Engagement (PPIE), community participation or other stakeholder advisory input was used to inform the design and implementation of the study, or to address equality, diversity and inclusion, share a description of their input. |
| g) Confirm if a brief protocol or proposal was developed that outlines the research/ evaluation questions, study design, methods of data collection, PPIE involvement, analysis plans, strategy to disseminate findings including potential audience, and provision of guidance on how to use data. If possible, share links to these documents. Report any changes made in the study protocol and the reason why these changes were made. |
| h) Share a description of the proposed duration of the study, and if any changes occurred, confirm the actual duration of the study including the data collection and data analysis periods. |
| i) Provide a clear description of the sampling approach, and the groups selected for the study, and explain why these approaches were taken. Clearly state if any groups or sites were not included in the study. |
| j) Adhere to good practices linked to informed consent, share a description of the process used for informed consent and recruitment of study participants. |
| **2. Evaluation or research team** |
| a) Provide a clear description and/or rationale of the team size (including any changes over time). |
| b) Describe the researcher's/ evaluator's relationship with (whether they have had previous engagement with the site) and in proximity to the research/ evaluation site. Including whether research/ evaluation is conducted virtually or face-to-face, or whether the researcher/evaluator is based in the area of the data collection. |
| c) Describe the levels of experience of team members and their backgrounds (including if any team members were part of the community, patient representatives or members of the public). |
| d) Indicate if team members received any training in rapid research/ evaluation methods. |
| e) Describe the roles and responsibilities of team members in this project and why the team was designed in this way. |
| f) If researchers/ evaluators reflected on how their background and experiences may have affected their data collection, analysis and interpretation, please describe this process (reflexivity). |
| **3. Data collection** |
| a) Clearly describe the data collection methods used throughout the study including any rapid methods, justify why these were selected and how they were implemented. |
| b) If there was any translation of materials, or if data was collected in another language, share the methods that were used to ensure that conceptual equivalence and cultural validity was achieved. |
| c) Provide information on any approaches, processes or practices used to ensure quality in data collection. |
| d) Provide information on any approaches, processes or practices used to ensure consistency in the methods of data collection across team members. |
| e) If data collection and analysis were carried out in parallel, describe the approaches, processes or practices used to facilitate this. |
| **4. Data analysis** |
| a) Clearly describe the methods that were used to analyse data. If different layers of analysis were carried out in parallel (i.e., rapid analysis and more in-depth analysis), describe the approaches, processes or practices used to facilitate this. |
| b) Provide information on any approaches, processes or practices used to ensure quality in data analysis. |
| c) Provide information on any approaches, processes or practices used to ensure consistency in the methods of data analysis across team members. |
| d) If relevant, provide a clear description of the type of data triangulation that was used and how triangulation was implemented. |
| e) Confirm if any findings were shared with stakeholders as the study was ongoing, report on what was shared, if feedback was received, and whether the feedback was used to make changes to the study design. |
| **5. Result interpretation** |
| a) Report if member checking was used (checking findings with study participants). Describe the approach that was used, how participant feedback was integrated, and, if not, describe why. |
| b) Describe how the findings from the study relate to the existing published literature. |
| c) If relevant to the study aims, report if there were any issues with the study design that prevented transferability or comparison to existing evidence and populations. |
| d) Confirm if any implications or recommendations were made based on the findings from the study. |
| e) Clearly report the limitations or gaps of the study. |
| **6. Dissemination** |
| a) Provide a clear description of the purpose and plan of dissemination and confirm if any changes occurred to the planned dissemination. |
| b) Describe whether dissemination was carried out as the study was ongoing and/or after the study ended. |
| c) Confirm if it is possible to access the raw data from the study on request. |
| **7. Impact** |
| a) If possible, report on how findings were used by the commissioners of the study and/or other stakeholders, and if they were not used as planned, share the reasons for this. |
| **8. Governance and accountability** |
| a) Include a statement on the regulatory and/or ethical approvals that were agreed, include any cases when these may not have been required and justify why. |
| b) Include a statement on the funding source. |
| c) Include a statement on any conflicts of interest. |

## 4. Discussion

### 4.1 A summary of the consensus-based study

A list of 38 standards were developed to enable the guidance, reporting and appraisal of rapid evaluation and appraisal studies. These standards were grouped into eight categories that could guide evaluators or researchers across different phases or aspects of the studies, including study design, the evaluation/research team dynamics; data collection; data analysis; result interpretation; dissemination; the impact of rapid studies; the governance and accountability of rapid studies.

A common issue raised across the consensus process was that understanding impact and dissemination of findings is not always possible, especially in the context of rapid evaluators who are commissioned by others to conduct the evaluation. It is often the commissioner who has insight into the dissemination and impact of the evaluation findings, and this is often not relayed back to the evaluation team. This is a challenge that has been discussed previously (Johnson & Vindrola-Padros, 2017) and is an area that needs to be addressed to enhance collaboration between commissioners and evaluators. A way to ensure this, could be through study protocols detailing dissemination plans and any impact measures that may be known by the commissioners. A similar comment was that some of the statements seemed more relevant in the context of rapid appraisals or rapid research when submitting papers for publication to an academic journal, rather than submission of rapid evaluations as internal reports to commissioners. Participants also flagged that some statements such as the researcher’s proximity to the study site, seemed more relevant for international contexts rather than domestic rapid evaluations. It will be important within the explanation and elaboration document to extend more on these statements, and flag that some statements may be more relevant to certain contexts. After testing STREAM in more contexts, it may become clearer if a shorter list of statements focused on specific contexts or study designs is required. This approach has been used previously as a result of feedback from Delphi studies (Staniszewska et al., 2017). This will be a factor that we will take into consideration in our plan to review and refine STREAM on a regular basis.

Some of the feedback received from the open comments in the first round of the study highlighted that participants thought the size or roles and responsibilities of team members should not determine the quality of a study. Some participants also shared in the open comments that they did not think training should be a necessity and some found it patronising. These statements related to teamwork will need to be justified more clearly in the explanation and elaboration document, especially if the statements are being used for critical appraisal. There will need to be specifications that if an evaluator/researcher shares in their study that a junior team member conducted the work, or that a small team conducted the work or that no members received training in rapid approaches, this does not mean that the study will rank poorly on the appraisal. Instead, emphasis should be placed on the fact that the study transparently listed the approaches that were used, adhering well to the standards.

### 4.2 Strengths and limitations

A key strength of this study is the diverse channels used to obtain feedback from stakeholders and experts in the field of rapid evaluation and appraisals. There were four opportunities to collect ample feedback – the steering group consultation, the three rounds of the e-Delphi, the stakeholder workshop and the piloting exercise. Across the opportunities for feedback and especially across the Delphi study, we were able to collect perspectives from stakeholders with diverse experiences in terms of their length of involvement with this field. In the Delphi study, this included a range of stakeholders with less than 5 years of experience who were likely to be involved in the day-to-day work of implementing new methods and could share insight into what would be useful in their practice. It also included those with more than 20 years of experience in the field, who may have a vast body of experience and understanding regarding how the field has changed.

Limitations of this study include the poor response rate to the e-Delphi invitation, of the 283 invited participants, only 16.6% participated in all three rounds of the Delphi. We assume the main reason for this low response rate is that those invited may not have had the capacity to participate. Of the participants that took part in the study, the majority (88.3%) of participants worked in the field of health. This is a limitation as within our eligibility criteria we had hoped to reach a broader audience that use rapid evaluation and appraisal methods. Similarly, the majority of Delphi respondents were based in high income countries, with 81.6% of participants based either in the UK or the USA, which meant that the views of stakeholders from countries with limited resources were limited. Both weaknesses limit the representativeness and generalisability of STREAM. Finally, we were only able to pilot STREAM in one context, within the reporting of a rapid evaluation conducted in the UK. This marks an area for the future development of STREAM and is discussed in the section below.

### 4.3 The future of STREAM

STREAM has been developed to improve the transparency in the reporting of rapid evaluations and appraisals. These standards will enable understanding in greater depth the methods that can be used across rapid studies to enhance their rigour and validity. The main purpose of STREAM will be to guide future evaluators and researchers in their study design, and to support them in reporting the approaches they used throughout their study when publishing findings in journals or submitting internal reports to commissioners. As STREAM will be published on the EQUATOR network it will be made accessible internationally to support the reporting of rapid evaluations and appraisals for publications in health-related journals. To strengthen STREAM further, the standards will need to be tested in more scenarios. This would include testing STREAM for use when setting up and implementing a study; piloting STREAM as a critical appraisal tool; using STREAM in different study designs such as rapid assessments; and using STREAM in different international contexts. Our research team has developed a plan to periodically review STREAM and test its usage across these contexts (S. E. Clark et al., 2022).

## 5. Conclusions

Rapid evaluations and appraisals can be useful in time and resource limited contexts and in the response to new or changing services, but close attention needs to be paid to their rigour and other factors that might influence the production of knowledge and validity of the findings. Generalising the use of STREAM by rapid evaluators, researchers and commissioners will address concerns around rigour, validity and transparency, while sharing findings in a timely way.

## Data Availability

All data produced in the present study are available upon reasonable request to the authors.

## Acknowledgements

We would like to thank the members of our steering group (Ginger Johnson; Patrick Nyikavaranda; Petra Gronholm; Stephanie Kumpunen); the participants of the e-Delphi study (those who wish to be named - Andrew Walker; Anna Dowrick; Ashley Hagaman; Claudia Canella; David Kryl; David Richard Brown; Georgia Chisnall; Heather Nunns; Itzel Eguiluz; Jackie Chandler; Jeremy Horwood; Judith Smith; Judy Margo; Karl McGrath; Kodjo Tehoungue; Krista Blair; Laura Holdsworth; Patricia Omidian; Patrick Nyikavaranda; Ruth Boyask; Sarah-Jane Fenton; Sergi Fàbregues); the stakeholders from the collaboration workshop (those who wish to be named - Alexandra Drought; Binta Sultan; Danielle Fairweather; Jackie Walumbe; Judith Yargawa; Julianna Smith; Kara Gray-Burrows; Paul McNaught; Sarah Williams; Yaz Osho); and the members of the wider Rapid Research Evaluation and Appraisal team that supported with piloting the Delphi survey, organising the workshop and piloting STREAM (Abinaya Chandrasekar; Becky Appleton; Germán Andrés Alarcón Garavito; Katie Gilchrist; Laura Maio; Noémie Déom; and Sophie Moniz).

