## Appendices for "Developing Standards for Rapid Evaluation and Appraisal Methods (STREAM): an e-Delphi consensus study"

Appendix A – Draft statements identified from the systematic review

| | |
|---|---|
| | **Study design** |
| 1. | Describe any theoretical frameworks or models used to guide the study design. |
| 2. | Describe any relevant reporting guidelines used throughout the study. |
| 3. | Describe any preliminary research or scoping studies carried out to inform the study design. |
| 4. | Define the purpose, aim or research questions guiding the study. |
| 5. | Indicate if stakeholders external to the evaluation or research team were involved in processes of co-design and how. |
| 6. | Confirm if a brief protocol or proposal was developed that outlines the research questions, study design, methods of data collection, and analysis and dissemination plans. If possible share links to these documents. Report any changes made in the study protocol and the reason why these changes were made. |
| 7. | Adhere to good practices linked to informed consent, share a description of the process used for informed consent and recruitment of study participants. |
| 8. | Provide a clear description of the sampling approach, and the groups selected for the study, and explain why these approaches were taken. Clearly state if any groups or sites were not included in the study due to time pressures. |
| | **Research team** |
| 1. | Provide a clear description of the research team size and why the team was designed in this way. |
| 2. | Describe the levels of experience of team members and their backgrounds. |
| 3. | Indicate if team members received any training within the timescale of the study. |
| 4. | Describe the roles and responsibilities of team members. |
| 5. | Provide information on any tools or techniques used to maintain consistency in data collection and analysis across team members. |
| | **Data collection** |
| 1. | Cleary describe the data collection methods used throughout the study, why these were selected and how they were implemented (including any changes made throughout the course of the study). |
| 2. | If time allows, pilot the methods, and describe the findings of the piloting activity (i.e., how these were used to make changes in the original methods). |
| 3. | If there was any translation of materials, or if data was collected in another language, share the methods that were used to ensure that conceptual equivalence and cultural validity was achieved. |
| 4. | Report on how quality and consistency was assured during data collection. |
| 5. | If data collection and analysis were carried out in parallel, describe the tools used to facilitate this process. |

| | |
|---|---|
| | **Data analysis** |
| 1. | Clearly describe the methods that were used to analyse data. If different layers of analysis were carried out in parallel (i.e., rapid analysis and more in-depth analysis), describe the tools used to facilitate these parallel processes. |
| 2. | Report on how quality and consistency was assured during data analysis. |
| 3. | If relevant, provide a clear description of the type of data triangulation that was used. |
| 4. | Confirm if any findings were shared with stakeholders as the study was ongoing, report on what was shared, if feedback was received, and whether the feedback was used to make changes to the study design. |
| | **Result interpretation** |
| 1. | Report if member checking was used. Describe the approach that was used, how participant feedback was integrated, and, if not, describe why. |
| 2. | Confirm if researchers reflected on their interpretations of the findings. |
| 3. | Describe how the findings from the study relate to the existing published literature. |
| 4. | Report if there were any issues with the study design that prevented the comparison to existing evidence and populations. |
| 5. | Clearly report the limitations of the study. |
| | **Dissemination** |
| 1. | Provide a clear description of the purpose of dissemination, reporting on whether the plans for dissemination were intended from the start or whether they changed throughout the study. |
| 2. | Clearly describe the plans for dissemination or the dissemination approach that was used. Describing whether dissemination was carried out as the study was ongoing and/or after the study ended. |
| 3. | Report on how findings were used, and if they were not used as planned, share the reasons for this. |
| | **Governance and accountability** |
| 1. | Include a statement on the regulatory approvals that were agreed, include any cases when these may not have been required and justify why. |
| 2. | If Patient and Public Involvement and Engagement or other stakeholder advisory input was used to inform the design and implementation of the study, include a description of their input. Include details of the stakeholders involved, the role they played, how collaboration was achieved and how their feedback was used. |

Appendix B – Statements for the first round of the Delphi survey (based on steering group feedback)

| | |
|---|---|
| | **Study design** |
| 1. | Describe any preliminary research, scoping studies or piloting methods to inform the study design. |
| 2. | Define the purpose, aim or research questions guiding the study. |
| 3. | Describe any theoretical frameworks or models used to guide the study design (including programme theories or theories of change in the case of rapid evaluations). |
| 4. | Provide a clear description of the intervention, programme or service being evaluated. |
| 5. | Describe any relevant reporting guidelines used throughout the study. |
| 6. | If Patient and Public Involvement and Engagement, community participation or other stakeholder advisory input was used to inform the design and implementation of the study, include a description of their input. Include details of the stakeholders involved, the role they played, how collaboration was achieved and how their feedback was used. |
| 7. | Confirm if a brief protocol or proposal was developed that outlines the research questions, study design, methods of data collection, analysis plans, strategy to disseminate findings, and provision of guidance on how to use data. If possible share links to these documents. Report any changes made in the study protocol and the reason why these changes were made. |
| 8. | Provide a clear description of the sampling approach, and the groups selected for the study, and explain why these approaches were taken. Clearly state if any groups or sites were not included in the study due to time pressures. |
| 9. | Adhere to good practices linked to informed consent, share a description of the process used for informed consent and recruitment of study participants. |
| | **Research team** |
| 10. | Provide a clear description of the research team size. |
| 11. | Share the geographic location of the researchers in connection to the research site. |
| 12. | Describe the levels of experience of team members and their backgrounds (including if any team members were part of the community, patient representatives or members of the public). |
| 13. | Indicate if team members received any training within the timescale of the study. |
| 14. | Describe the roles and responsibilities of team members in this project and why the team was designed in this way. |
| | **Data collection** |
| 15. | Cleary describe the data collection methods used throughout the study, why these were selected and how they were implemented. |
| 16. | If there was any translation of materials, or if data was collected in another language, share the methods that were used to ensure that conceptual equivalence and cultural validity was achieved. |

| | |
|---|---|
| 17. | Provide information on any tools or techniques used to ensure quality and maintain consistency in data collection across team members. |
| 18. | If data collection and analysis were carried out in parallel, describe the tools used to facilitate this process. |
| | **Data analysis** |
| 19. | Clearly describe the methods that were used to analyse data. If different layers of analysis were carried out in parallel (i.e., rapid analysis and more in-depth analysis), describe the tools used to facilitate these parallel processes. |
| 20. | Provide information on any tools or techniques used to ensure quality and maintain consistency in data analysis across team members. |
| 21. | If relevant, provide a clear description of the type of data triangulation that was used. |
| 22. | Confirm if any findings were shared with stakeholders as the study was ongoing, report on what was shared, if feedback was received, and whether the feedback was used to make changes to the study design. |
| | **Result interpretation** |
| 23. | Report if member checking was used. Describe the approach that was used, how participant feedback was integrated, and, if not, describe why. |
| 24. | Confirm if researchers reflected on their interpretations of the findings, in terms of how the roles and backgrounds of different team members may have impacted their interpretations. |
| 25. | Describe how the findings from the study relate to the existing published literature. |
| 26. | Report if there were any issues with the study design that prevented the comparison to existing evidence and populations. |
| 27. | Confirm if any implications or recommendations were identified from the results of the study. |
| 28. | Clearly report the limitations of the study. |
| | **Dissemination** |
| 29. | Provide a clear description of the purpose of dissemination. |
| 30. | Report on whether the plans for dissemination were intended from the start or whether they changed throughout the study. |
| 31. | Clearly describe the plans for dissemination or the dissemination approach that was used. |
| 32. | Describe whether dissemination was carried out as the study was ongoing and/or after the study ended. |
| 33. | If possible, report on how findings were used by the commissioners of the study and/or other stakeholders, and if they were not used as planned, share the reasons for this. |
| | **Governance and accountability** |
| 34. | Include a statement on the regulatory approvals that were agreed, include any cases when these may not have been required and justify why. |
| 35. | Include a statement on the funding source. |
| 36. | Include a statement on any conflicts of interest. |

Appendix C – Statements for the second round of the Delphi survey (based on open feedback from the first round of the Delphi)

| | |
|---|---|
| | **Study design** |
| 1. | Define the purpose, aim or research questions and planned deliverables guiding the study. |
| 2. | Describe any preliminary research, scoping studies or piloting methods to inform the study design. |
| 3. | Describe any theoretical frameworks or models used to guide the study design (including programme theories or theories of change in the case of rapid evaluations). |
| 4. | Provide a clear description of the intervention, programme or service being evaluated. |
| 5. | Indicate any relevant reporting guidelines used throughout the study. |
| 6. | If Patient and Public Involvement and Engagement (PPIE), community participation or other stakeholder advisory input was used to inform the design and implementation of the study, or to address equality, diversity and inclusion, share a description of their input. |
| 7. | Confirm if a brief protocol or proposal was developed that outlines the research questions, study design, methods of data collection, PPIE involvement, analysis plans, strategy to disseminate findings, and provision of guidance on how to use data. If possible share links to these documents. Report any changes made in the study protocol and the reason why these changes were made. |
| 8. | Share a description of the proposed duration of the study, and if any changes occurred, confirm the actual duration of the study including the data collection and data analysis periods. |
| 9. | Provide a clear description of the sampling approach, and the groups selected for the study, and explain why these approaches were taken. Clearly state if any groups or sites were not included in the study due to time pressures. |
| 10. | Adhere to good practices linked to informed consent, share a description of the process used for informed consent and recruitment of study participants. |
| | **Research team** |
| 11. | Provide a clear description of the research team size (including any changes over time). |
| 12. | Describe the researcher's relationship with and in proximity to the research site. Including whether research is conducted virtually or face-to-face, or whether the researcher is based in the area of the data collection. |
| 13. | Describe the levels of experience of team members and their backgrounds (including if any team members were part of the community, patient representatives or members of the public). |
| 14. | Indicate if team members received any training in rapid research methods. |
| 15. | Describe the roles and responsibilities of team members in this project and why the team was designed in this way. |
| 16. | If researchers reflected on how their background and experiences may have affected their data collection, analysis and interpretation, please describe this process. |

| | |
|---|---|
| | **Data collection** |
| 17. | Cleary describe the data collection methods used throughout the study including any rapid methods, justify why these were selected and how they were implemented. |
| 18. | If there was any translation of materials, or if data was collected in another language, share the methods that were used to ensure that conceptual equivalence and cultural validity was achieved. |
| 19. | Provide information on any approaches, processes or practices used to ensure quality and to use methods in data collection consistently across team members. |
| 20. | If data collection and analysis were carried out in parallel, describe the approaches, processes or practices used to facilitate this. |
| | **Data analysis** |
| 21. | Clearly describe the methods that were used to analyse data. If different layers of analysis were carried out in parallel (i.e., rapid analysis and more in-depth analysis), describe the approaches, processes or practices used to facilitate this. |
| 22. | Provide information on any approaches, processes or practices used to ensure quality and to use methods in data analysis consistently across team members. |
| 23. | If relevant, provide a clear description of the type of data triangulation that was used and how triangulation was implemented. |
| 24. | Confirm if any findings were shared with stakeholders as the study was ongoing, report on what was shared, if feedback was received, and whether the feedback was used to make changes to the study design. |
| | **Result interpretation** |
| 25. | Report if member checking was used (checking findings with participants). Describe the approach that was used, how participant feedback was integrated, and, if not, describe why. |
| 26. | Describe how the findings from the study relate to the existing published literature. |
| 27. | If relevant to the study aims, report if there were any issues with the study design that prevented transferability, generalisability or comparison to existing evidence and populations. |
| 28. | Confirm if any implications or recommendations were made based on the findings from the study. |
| 29. | Clearly report the limitations of the study. |
| | **Dissemination** |
| 30. | Provide a clear description of the purpose and plan of dissemination, and confirm if any changes occurred to the planned dissemination. |
| 31. | Describe whether dissemination was carried out as the study was ongoing and/or after the study ended. |
| 32. | If possible, report on how findings were used by the commissioners of the study and/or other stakeholders, and if they were not used as planned, share the reasons for this. |
| 33. | Confirm if it is possible to access the raw data from the study on request. |
| | **Governance and accountability** |

| |
|---|
| 34. Include a statement on the regulatory and/or ethical approvals that were agreed, include any cases when these may not have been required and justify why. |
| 35. Include a statement on the funding source. |
| 36. Include a statement on any conflicts of interest. |
